# Spectral Changes in Motor Thalamus Field Potentials during Movement

**DOI:** 10.1101/2024.09.11.24313483

**Authors:** Bryan T. Klassen, Matthew R. Baker, Michael A Jensen, Gabriela Ojeda Valencia, Kai J. Miller

**Author notes:** Correspondence: Bryan T. Klassen.

## Abstract

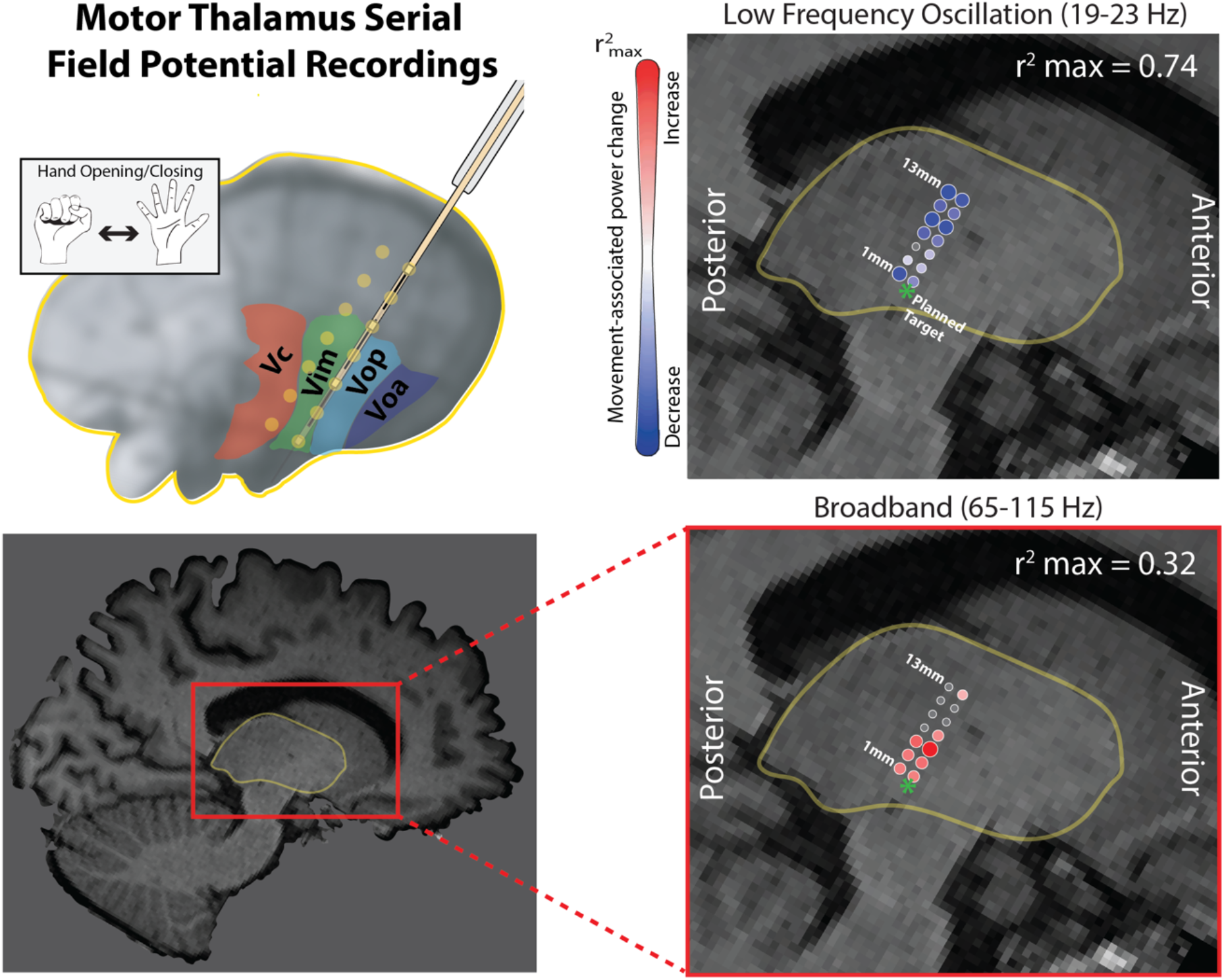

The motor thalamus plays a crucial role in the integration and modulation of sensorimotor information and projects to the primary motor cortex. While voltage power spectral changes in the motor cortex with movement have been well-characterized, the corresponding activity in the motor thalamus, particularly broadband (sometimes referred to as high gamma) power, remains unclear. The present study aims to characterize spectral changes in the motor thalamus during hand movements of 15 subjects undergoing awake deep brain stimulation surgery targeting the ventral intermediate (VIM) nucleus of the thalamus for disabling tremor. We analyzed power changes in subject-specific low frequency oscillations (<30 Hz) and broadband power (captured in 65-115 Hz band) of serial field potential recordings. Consistent with previous studies, we found widespread decreases in low-frequency oscillations with movement. Importantly, in most subjects we also observed a significant increase in broadband power, primarily in the inferior recording sites corresponding with estimated VIM region. One subject also performed an imagined movement task during which low frequency oscillatory power was suppressed. These electrophysiologic changes may be leveraged as biomarkers for thalamic functional mapping, DBS targeting, and closed loop applications.

**NEW & NOTEWORTHY:** We studied movement-associated spectral changes in the human motor thalamus of subjects undergoing awake deep brain stimulation surgery. We observed focal increases in broadband power in the motor thalamus with movement. This biomarker may be used as a tool for intraoperative functional mapping, DBS targeting, and closed-loop device control.

## INTRODUCTION

The thalamus plays a crucial role in the modulation and integration of sensorimotor information(1, 2). The motor thalamus is an intricate structure with functional topography parcellated by the motor homunculus and has overlapping representations from basal ganglia and cerebellar nuclei, with major projections to primary motor cortex (M1)(3-5). Motor-related cortical spectral changes have been well-described in human electrocorticography (ECoG) and stereoencephalography (sEEG) studies, revealing spatially distributed low-frequency (<30Hz) power suppression accompanied by focal broadband (sometimes referred to as high gamma) power increases for both overt and imagined movements(6-10). Previous studies have associated lower frequency band power with thalamocortical circuits, and broadband changes with very local increases in neuron population firing rates(11-13). Like cortical activity, motor thalamus movement-related oscillatory activity in low-frequency bands is suppressed during movement(14). However, broadband power changes have not been explored.

The present study aims to characterize movement-related spectral changes in the motor thalamus. The posterior region of the motor thalamus, the ventral intermedius (VIM) nucleus, relays motor signals between the cerebellum and M1 and is a key therapeutic target for deep brain stimulation (DBS) for tremor and other movement disorders(15). This therapeutic targeting provides an opportunity to study motor-related spectral changes in subcortical structures with direct input to M1. Specifically, serial field potential recordings from the VIM nucleus in awake subjects undergoing DBS lead placement surgery for tremor were analyzed for hand movement-associated electrical potential power spectral changes in 15 subjects. One subject also performed an imagined movement task.

Consistent with previous studies, we find that in low frequency ranges (<30Hz), distinct spectral peaks are present in rest intervals, and decrease in power during movement periods with a spatially broad distribution across the motor thalamus(**Fig. 1)**(14) At higher frequencies (captured by 65-115 Hz band), we observed movement-associated increases in power most often in the inferior recording sites for most subjects, near the predicted VIM nucleus. Interestingly, low frequency power decreases were observed in both overt and imagined movement, whereas broadband power increases were only seen in overt movement.

**Figure 1.**
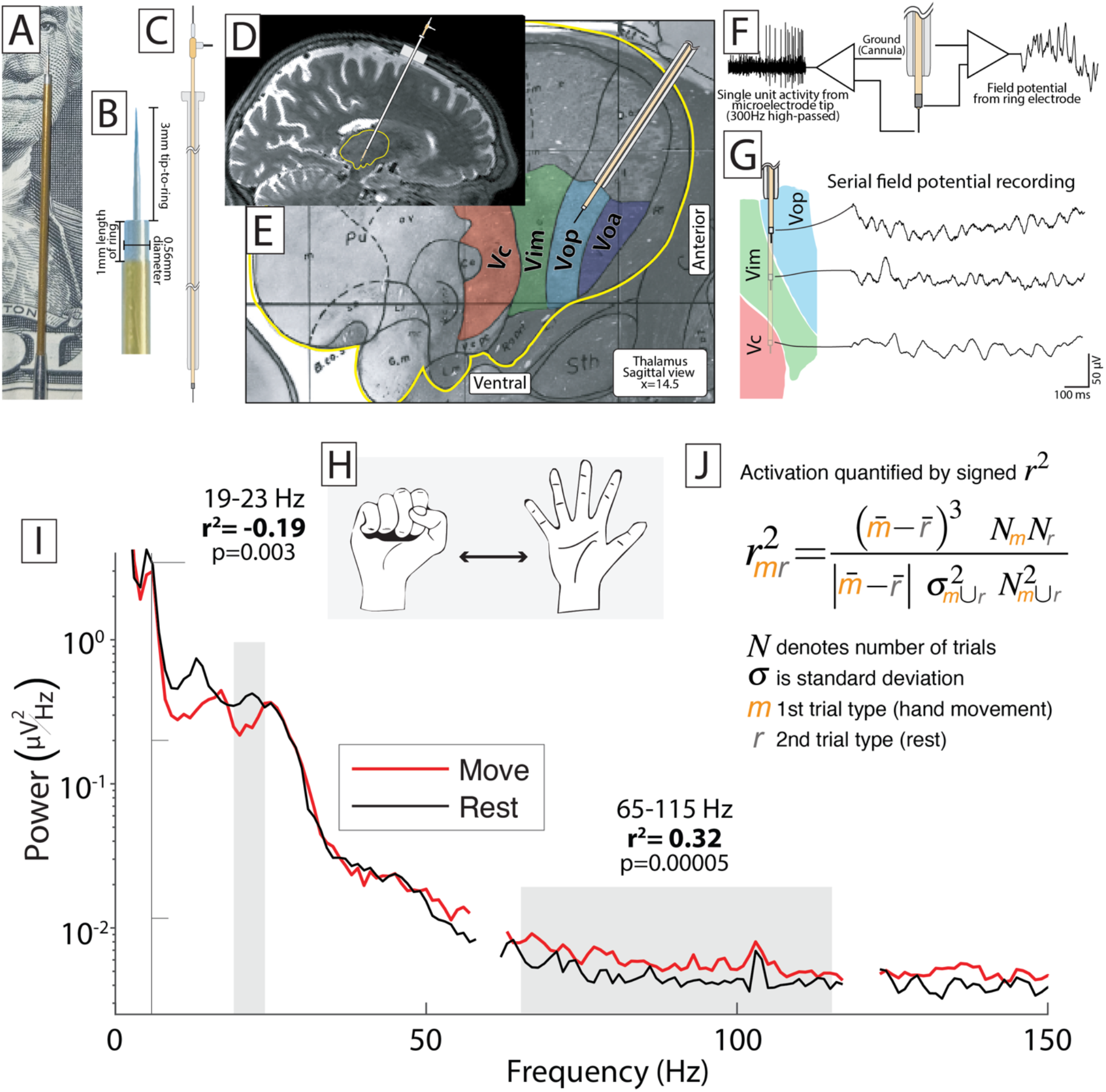
Experimental and Signal Processing Methods. (**A**) Recordings were obtained using a clinical DBS electrode both (B) with a high-impedance tip and with a low-impedance ring having surface area of 0.56 mm^2^. The electrode was passed through a cannula (**C**) that had been inserted into the dorsal thalamus, as shown on a sagittal MRI slice (**D**), aligned along a trajectory targeting the VIM thalamic nucleus (**E**). (**F**) Field potentials were recorded from the ring electrode and single unit activity was recorded from the microelectrode tip; the shaft of the cannula was referenced in all cases. (**G**) This study analyzes the field potentials at multiple evenly spaced sites along the trajectory recorded serially as the electrode was passed to target. **(H)** at each recording site, subjects performed a simple motor task alternating between opening/closing of the hand and rest. **(I)** Averaged power spectral densities (PSD) from one recording site for Subject 1 are shown for movement and rest epochs. In this case, power within low frequency (<30 Hz) oscillations decreased during movement while broadband spectral power (65-115 Hz) increased. PSD for a subject were normalized to the global mean across all trials, and a signed r^2^ cross-correlation value was calculated from the mean PSD for movement versus rest trials, as shown in equation **(J)**. The r^2^ for both broadband change and change in a subject-specific 5 Hz wide low frequency band were computed.

## MATERIALS AND METHODS

### Subjects

Fifteen patients undergoing deep brain stimulation electrode placement into the bilateral ventralis intermedius (Vim) nucleus of the thalamus for treatment of disabling tremor consented to participate as subjects in a research protocol during the awake surgery. The study and consent procedures were approved by Mayo Clinic’s internal review board (IRB #19-009878).

### Recordings

Serial thalamic recordings during task performance were obtained at multiple evenly-spaced recording sites (2 or 3 mm apart) as a hybrid microelectrode/macroelectrode (AlphaOmega Sonus STR-009080-00) was advanced towards the inferior border of the thalamus (**Fig. 1A-G**). In 4 subjects, simultaneous recordings were obtained from two electrodes arranged parallel to each other along the anterior-posterior plane. Data were recorded to an AlphaOmega Neuromega system, referenced to the shaft of the electrode cannula with sampling rate of 44 kHz. Surface EMG was recorded using pairs of bipolar-referenced Ag/AgCl electrodes placed 2 cm apart overlying the forearm muscles for finger flexion/extension.

### Motor Task

At each recording site, subjects were verbally and visually cued for two alternating block conditions: 1) continuous opening/closing movements of the dominant hand, 2) rest. Individual task epochs were approximately 5 seconds in duration. The move-rest sequence was repeated 20 times per site. Compliance with the task was assessed in real-time by observing subject, movement and monitoring EMG activity. The imagined movement task was performed in the same manner, with “imagined movement” periods being cued verbally by the experimenter.

### Signal Processing

All analyses were performed in MATLAB. Trial epochs of movement versus rest were manually segmented via visual inspection of the rectified EMG; ambiguous epochs were rejected. Averaged power spectral densities (PSD) for individual movement or rest epochs were calculated from 1 to 300 Hz, with 1 Hz frequency resolution, using Welch’s method of overlapping periodograms with a 1 second Hann window and 0.5 second overlap to attenuate edge effects. Averaged PSDs for each trial were then normalized to the global mean across all trials. At each recording site, we calculated signed r^2^ cross-correlation values (r^2^) by comparing the mean PSD between movement and rest trials (**Fig. 1H-J**). As a proxy for broadband activity, r^2^ was calculated for the 65-115 Hz band (avoiding line noise at 60 and 120 Hz). In cortex, broadband changes reflect the 1/f shape of the power spectrum and correlate with activity in local neuronal populations **(Fig. 1)**(11). Because the center frequency of movement-related oscillatory shifts in the lower (<30 Hz) frequency bands varied between subjects, we individually determined the center frequency for each subject (and separately for overt versus imagined movement in subject 15). We identified 5 consecutive frequency bins within the 8-30 Hz range that were able to maximally discriminate between movement and rest as follows: 1) r^2^ values for each 1 Hz bin within the 8-30 Hz range were averaged across all recording sites in a subject and 2) the 5 contiguous bins with the highest sum of r^2^ values was selected. The lower frequency bands have been associated with thalamocortical circuits(16).

### Plotting Power Changes on Subject-Specific MRI

The site of each recording relative to the final lead position was documented intraoperatively and could be computed using offsets to the lead artifact as seen on a postoperative CT. The CT was then co-registered to a preoperative T1-weighted MRI using mutual information in SPM12(17). The MRI was resliced in plane with recording sites and served as the background on which data were plotted. Low frequency band and broadband plots were prepared for movement associated power. We set significance at p<0.05, uncorrected. Recording sites with a significant r^2^ were plotted in red (movement associated increase) or blue (movement associated decrease), with the sizes scaled to the maximum r^2^ value for that subject (**Fig. 2**). Sites without significant power change were plotted with an open circle of fixed diameter.

**Figure 2:**
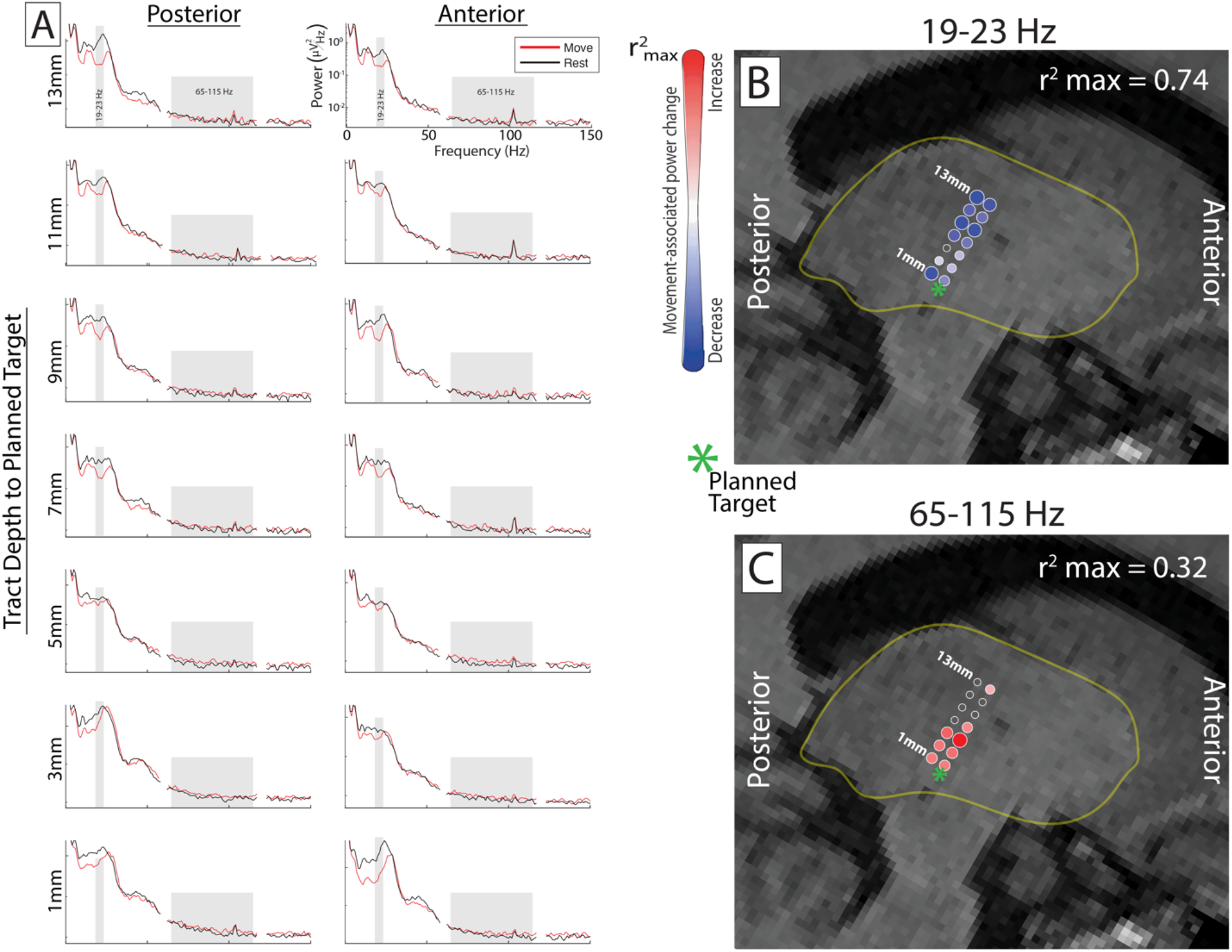
Spectra and r^2^ plots for a single subject. (**A**) Average power spectral densities during movement (red) and rest (black) shown for each of 14 recording sites sampled in Subject 1. The low (19-23 Hz) and broadband (65-115) frequency ranges are highlighted with gray boxes. The r^2^ values at each site are shown for (**B**) the 19-23 Hz band and (**C**) the 65-115 Hz band as dots plotted on sagittal T1 MRI slice. A red dot indicates an increase in power with movement, and a blue dot indicates a decrease. The size of each dot represents the absolute value of r^2^ scaled to the maximum r^2^ value for the frequency band (as shown in upper right-hand corner of each plot). In this subject, the sites were arranged along two parallel tracts 2mm apart in the anterior/posterior plane with serial recordings taken every 2 mm during electrode descent to the planned target (green asterisk).

## RESULTS

### Low Frequency Oscillations

Example power spectral densities (PSD) for movement and rest epochs from one recording site for Subject 1 are shown in **figure 1I. Figure 2** shows the spectra at each recording site for this single subject (**A**) and r^2^ maps for the low frequency band (**B**) and broadband (**C**).

We found that the low frequency band showing the greatest power change with movement varied between subjects, from as low as 8-12 Hz, up to 20-24 Hz. Significant decreases in power within the subject-specific low frequency bands were present in at least one recording site for 12 of 15 total subjects (r^2^_max_ range = 0.39-0.74)(**Fig. 3C**). In two cases (Subjects 6 and 7) there was increased power within the band, and in one (case 8) there was no significant change with movement. Sites showing a significant power decrease in the low frequency band were widely distributed through the sampled region of dorsal and ventral thalamus.

**Figure 3:**
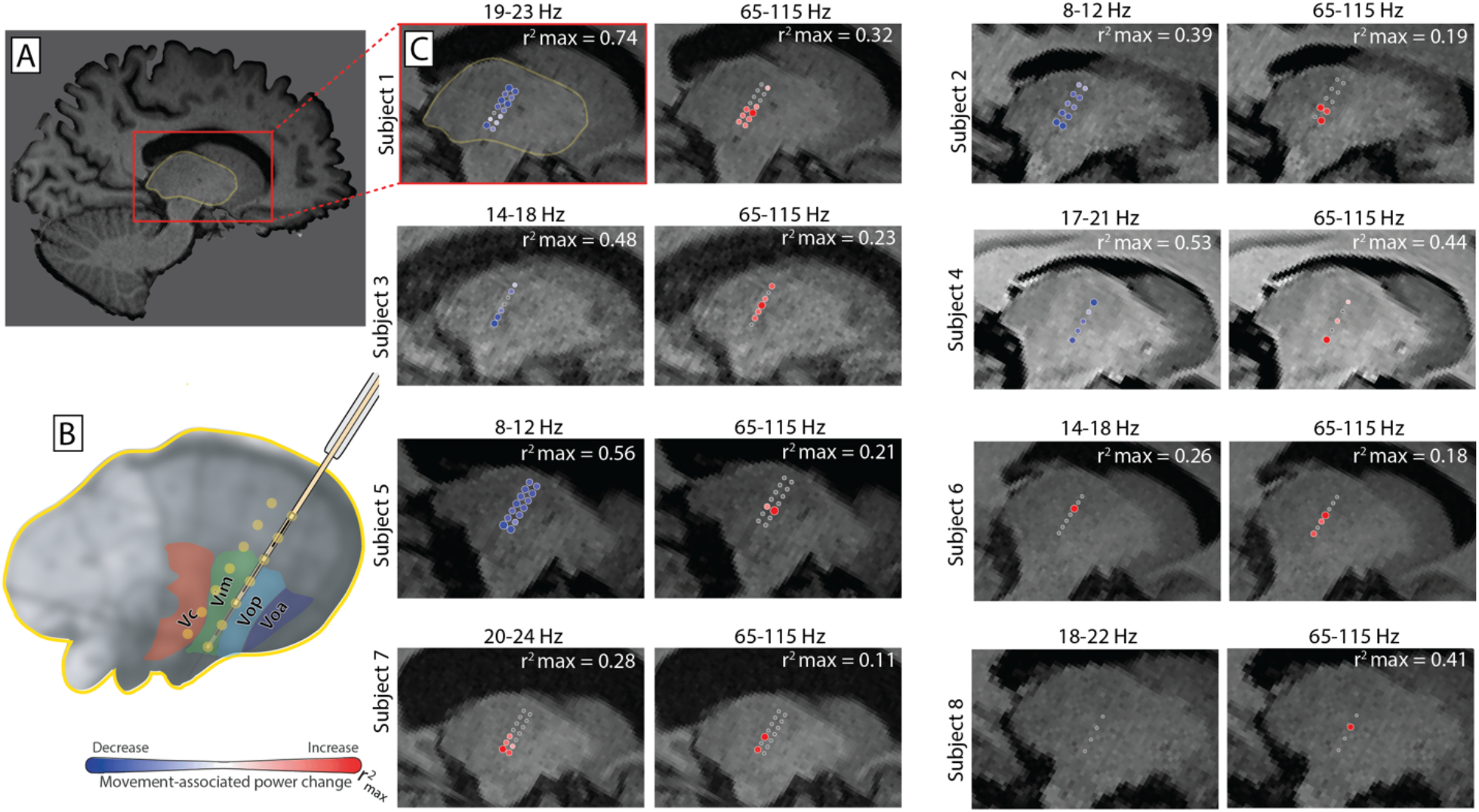
Subjects with significant increase in broadband spectral power during movement. (**A**) A sagittal T1 MRI slice from Subject 1, showing the thalamus (yellow outline) and the cropped/enlarged region used for r^2^ plots (red box). (**B**) Cartoon thalamus showing the ventral sensorimotor nuclei including the VIM target. Approximate recording locations are shown relative to these nuclei for the case of two parallel tracts (anterior/posterior) and for seven recording sites spaced 2mm apart. (**C**) All subjects with significant movement associated broadband increase. The r^2^ at each recording site is plotted on a sagittal T1 MRI for both the subject-specific low frequency band and broadband. Red dots indicate and increase in spectral power with movement, blue a decrease. The size of each dot represents the absolute value of r^2^, scaled to the maximum r^2^ value for that band.

### Broadband Spectral Changes

A significant increase in broadband power during movement was observed in at least one recording site for 9 of the 15 subjects (r^2^_max_ range = 0.11-0.44) (**Fig. 3C**). Sites showing significant broadband power increase were most often found at the inferior recording sites, in the ventral thalamus near the predicted region of the VIM nucleus.

### Overt versus Imagined Movement

In addition to the normal hand opening/closing task, Subject 15 performed imagined hand movements. “Imagined movement” periods were cued verbally by the experimenter. EMG confirmed that no active movements contaminated imagined movement epochs (**Fig. 4A-B**). Low frequency oscillations were evaluated at 22-26 Hz for movement, and 19-23 Hz for imagined movement (**Fig 4C**). We observed significant power decreases in the low frequency band at both recording sites for overt (r^2^_max_ = 0.48) and imagined (r^2^_max_ = 0.23) movements (**Fig 4D**). For overt movement, there was a significant increase in broadband power at the dorsal site only (r^2^ = 0.33). For imagined movement there were no significant changes in broadband power.

**Figure 4.**
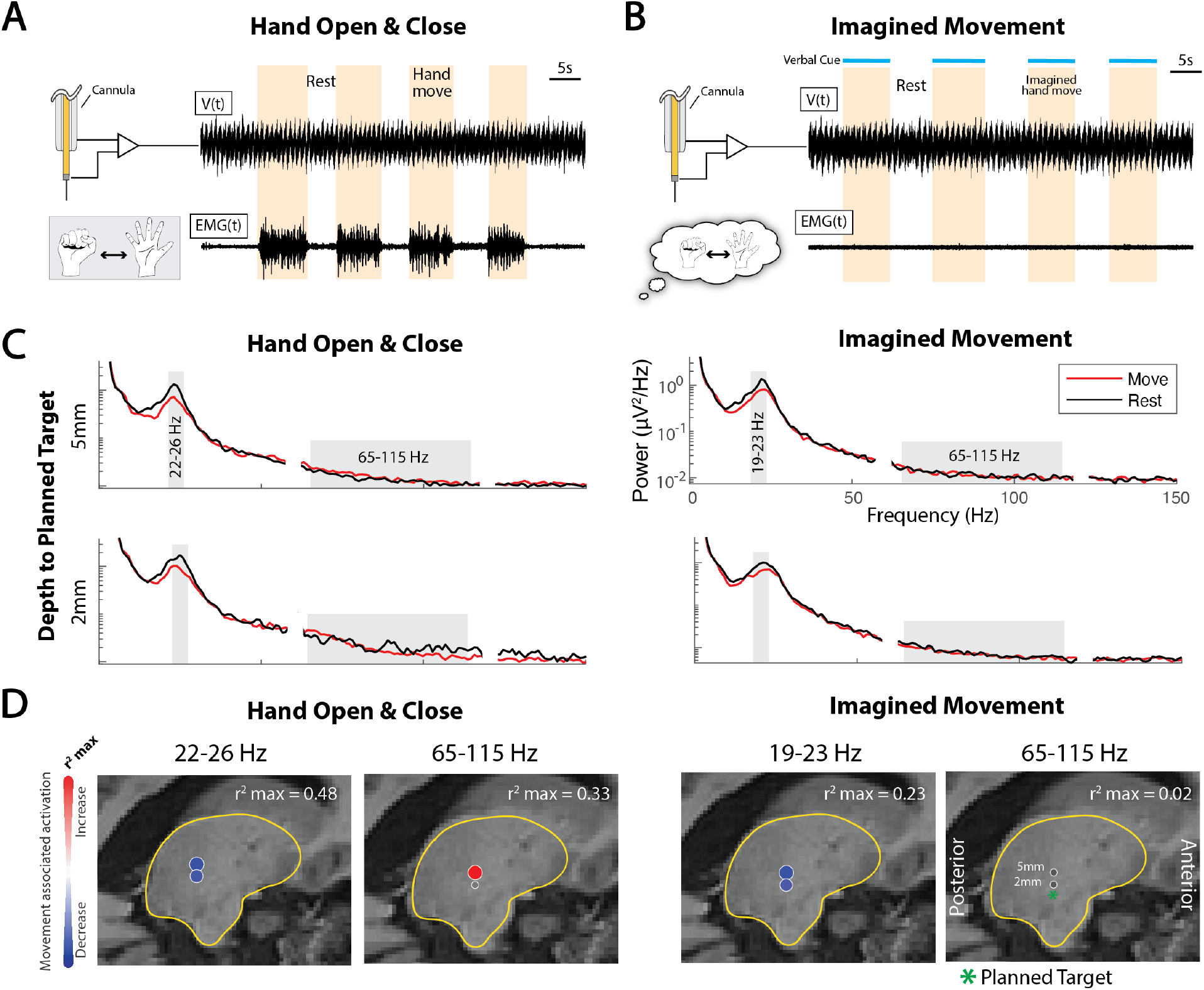
Spectra and r^2^ plots for overt and imagined hand movement. **(A)** Macroelectrode recording from the VIM nucleus of the thalamus during overt hand movement (shown on EMG recording) in subject 15. **(B)** VIM Ring electrode and EMG recordings during imagined hand movement. **(C)** Averaged power spectral densities (PSD) from two VIM recording sites during overt and imagined hand movement. Significant low frequency band (LFB; <30 Hz) suppression was observed at both sites during both tasks, whereas broadband power (65-115 Hz) increased at the 5mm above target site, but not during imagined movement. **(D)** The LFB and broadband frequency r^2^ values plotted on sagittal T1 MRI slice.

## DISCUSSION

Although they have limited spatial resolution, single-unit microelectrode recordings in deep brain nuclei are commonly used to identify specific brain regions and guide placement of deep brain stimulating electrodes (**Fig. 1A-G**)(18). Macroelectrode recordings, in contrast, reflect the summated activity of many neuronal events over a wider region surrounding the electrode. At this scale, power shifts in distinct spectral peaks (oscillations/rhythms) correspond to large-scale coherence within a specific timescale and frequency range. In contrast, asynchronous focal increases in electrical activity are revealed by broad spectral increases in a 1/f shape of the power spectrum (broadband) and have been shown to reflect the summated rate of neuronal firing near the electrode(**Fig 1I-J**)(11, 19). Surface macroelectrode recordings (ECoG) from human primary motor cortex show that voluntary movement leads to desynchronization of oscillations in the beta frequency range, which is spatially diffuse with significant overlap in regions suppressed regardless of which body segment is moved(6, 20). Conversely, movement-associated increases in broadband spectral power are spatially discrete enough to allow for discrimination between movement of different fingers in the same hand(21).

The reciprocal connections between cortical motor regions and subcortical nuclei, including the motor relay nuclei within the ventral thalamus, play a key role in generation and coordination of voluntary movement (**Fig. 1D**). Information processed in the basal ganglia is passed, via the *ventralis oralis* anterior (VOA) and posterior (VOP) thalamic nuclei, to higher-order cortical motor regions (supplementary and pre-motor areas), playing a key role in modulating the initiation and scaling of internally generated movement, whereas the VIM thalamic nuclei direct outputs from the cerebellar nuclei to primary motor cortex, correcting movements to match the environment(3, 5, 16). Desynchronization of beta frequency range oscillations has been observed in human motor thalamus during voluntary movement(14). High frequency gamma oscillation peaks (55-80 Hz) in human motor thalamus have been observed in a go no-go task(22); however, movement-associated broadband increases have not been described previously.

In the present study we observed broadband power increases in most subjects during a simple verbal and visually-cued voluntary hand movement task. For subjects in whom this was observed, sites showing significant broadband power increases tended to be more spatially discrete (near ventral/inferior thalamus, presumably VIM) than were sites showing desynchronization in low frequency oscillations (**Fig 2-3**), complementing what has previously been observed in motor cortex(6). These findings could serve as a new means to functionally map the motor thalamus during surgical placement of thalamic DBS electrodes for treatment of tremor. This is particularly important since motor thalamus subregion boundaries are not visible on standard MRI series, and single-unit activity recordings are inherently spatially limited.

These power spectral changes might be utilized as a control signal biomarker for closed-loop DBS paradigms. Chronic VIM stimulation can be associated with ataxic side-effects including speech disturbance, limb incoordination, or gait disturbance, and a subset of subjects develop tolerance with chronic stimulation(23, 24). Thus, there is interest in approaches that would apply stimulation only as needed to control movement and reduce or withdraw stimulation during rest. Early work has focused on using ECoG feedback for control(10, 14, 25, 26); however, this requires implantation of additional electrodes. Therefore, leveraging thalamic LFP recorded from the stimulating electrode itself is appealing, and could be used for patients that already have implants, sparing them from further cranial surgery. Early approaches along these lines have used low frequency oscillation desynchronization as a control signal(14, 27). Our data suggest that broadband power shifts may also be used. The increased somatotopic resolution made possible by the focal nature of broadband shifts, as well as the differential effects of overt movement (decreased oscillations, increased broadband) and imagined movement (decreased oscillations, no change in broadband)(**Fig 4**) may allow decoding of higher degrees of freedom to use as control signals for other applications including brain computer interface.

In most subjects, a diffuse desynchronization of low frequency oscillations was seen during movement. Qualitative review of the spectra shows that there is often more than one oscillation in this range, and that in some cases the most prominent oscillation shifts its center frequency with movement. We focused our analysis to the 5 Hz band yielding the highest r^2^ value to identify subject-specific low frequency oscillatory changes. While this was useful to compare overall changes in oscillatory activity to the broadband power shifts, future studies may consider multiple distinct oscillations in the 8 to 30 hertz range.

We did not observe movement-associated increase in broadband power in all subjects. This may be because the frequency range of interest approaches the noise floor of our recording paradigm. The operating room has many idiosyncratic sources of electrical noise which vary from case-to-case and within a single surgical case, potentially obscuring subtle power shifts. It is also possible that the serial nature of recordings did not represent exact identical behavioral states (attention, etc.). Future studies will focus on simultaneous recordings at multiple sites, multiple movement types (i.e. hand, foot, tongue), and using higher-grade amplifiers to overcome these limitations.

Overall, our findings advance the understanding of the motor thalamus’s role in movement, and may be relevant to tremor mechanisms, intraoperative functional mapping, DBS targeting, and closed-loop approaches for essential tremor and other movement disorders.

## Data Availability

All data necessary to interpret, verify, and extend the research will be anonymized and made publicly available with the final version of this article. A README document will be included detailing data variables. Code used to perform analyses and reproduce the illustrations will also be included.

## DATA AVAILABILITY

All data recorded necessary to interpret, verify, and extend the research will be anonymized and made publicly available with the final version of this article. A README document will be included, which details all variables included in the data files. All code used to perform analyses and reproduce the illustrations will also be publicly available.

## ACKNOWLEDGMENTS

We are grateful to the patients who volunteered their time to participate in this research and the clinical staff at St. Mary’s Hospital.

## AUTHOR CONTRIBUTIONS

BTK and KJM conceived experiments. BTK, MRB, and KJM collected the data and performed the experiments. BTK, MRB, GOV, and MAJ performed the data curation. BTK, MRB, and KJM analyzed the data. The original draft was written by BTK, MRB, and KJM and was revised by and edited by all authors. BTK and KJM supervised the project.

## REFERENCES

1. Cappe C, Morel A, Barone P, and Rouiller EM. The thalamocortical projection systems in primate: an anatomical support for multisensory and sensorimotor interplay. Cereb Cortex 19: 2025–2037, 2009.

2. Shine JM, Lewis LD, Garrett DD, and Hwang K. The impact of the human thalamus on brain-wide information processing. Nat Rev Neurosci 24: 416–430, 2023.

3. Bosch-Bouju C, Hyland BI, and Parr-Brownlie LC. Motor thalamus integration of cortical, cerebellar and basal ganglia information: implications for normal and parkinsonian conditions. Front Comput Neurosci 7: 163, 2013.

4. Vitek JL, Ashe J, DeLong MR, and Alexander GE. Physiologic properties and somatotopic organization of the primate motor thalamus. J Neurophysiol 71: 1498–1513, 1994.

5. Vitek JL, Ashe J, DeLong MR, and Kaneoke Y. Microstimulation of primate motor thalamus: somatotopic organization and differential distribution of evoked motor responses among subnuclei. J Neurophysiol 75: 2486–2495, 1996.

6. Miller KJ, Leuthardt EC, Schalk G, Rao RP, Anderson NR, Moran DW, Miller JW, and Ojemann JG. Spectral changes in cortical surface potentials during motor movement. J Neurosci 27: 2424–2432, 2007.

7. Jensen MA, Huang H, Valencia GO, Klassen BT, van den Boom MA, Kaufmann TJ, Schalk G, Brunner P, Worrell GA, Hermes D, and Miller KJ. A motor association area in the depths of the central sulcus. Nat Neurosci 26: 1165–1169, 2023.

8. Levine SP, Huggins JE, BeMent SL, Kushwaha RK, Schuh LA, Passaro EA, Rohde MM, and Ross DA. Identification of electrocorticogram patterns as the basis for a direct brain interface. J Clin Neurophysiol 16: 439–447, 1999.

9. Crone NE, Miglioretti DL, Gordon B, Sieracki JM, Wilson MT, Uematsu S, and Lesser RP. Functional mapping of human sensorimotor cortex with electrocorticographic spectral analysis. I. Alpha and beta event-related desynchronization. Brain 121 (Pt 12): 2271–2299, 1998.

10. Leuthardt EC, Schalk G, Wolpaw JR, Ojemann JG, and Moran DW. A brain-computer interface using electrocorticographic signals in humans. J Neural Eng 1: 63–71, 2004.

11. Manning JR, Jacobs J, Fried I, and Kahana MJ. Broadband shifts in local field potential power spectra are correlated with single-neuron spiking in humans. J Neurosci 29: 13613–13620, 2009.

12. Miller KJ, Sorensen LB, Ojemann JG, and den Nijs M. Power-law scaling in the brain surface electric potential. PLoS Comput Biol 5: e1000609, 2009.

13. Miller KJ, Hermes D, Honey CJ, Sharma M, Rao RP, den Nijs M, Fetz EE, Sejnowski TJ, Hebb AO, Ojemann JG, Makeig S, and Leuthardt EC. Dynamic modulation of local population activity by rhythm phase in human occipital cortex during a visual search task. Front Hum Neurosci 4: 197, 2010.

14. Guehl D, Guillaud E, Langbour N, Doat E, Auzou N, Courtin E, Branchard O, Engelhardt J, Benazzouz A, Eusebio A, Cuny E, and Burbaud P. Usefulness of thalamic beta activity for closed-loop therapy in essential tremor. Sci Rep 13: 22332, 2023.

15. Shanker V. Essential tremor: diagnosis and management. BMJ 366: l4485, 2019.

16. Opri E, Cernera S, Okun MS, Foote KD, and Gunduz A. The Functional Role of Thalamocortical Coupling in the Human Motor Network. J Neurosci 39: 8124–8134, 2019.

17. Richner TJ, Klassen BT, and Miller KJ. An in-plane, mirror-symmetric visualization tool for deep brain stimulation electrodes. Annu Int Conf IEEE Eng Med Biol Soc 2020: 1112–1115, 2020.

18. Vinke RS, Geerlings M, Selvaraj AK, Georgiev D, Bloem BR, Esselink RAJ, and Bartels R. The Role of Microelectrode Recording in Deep Brain Stimulation Surgery for Parkinson’s Disease: A Systematic Review and Meta-Analysis. J Parkinsons Dis 12: 2059–2069, 2022.

19. Miller KJ, Honey CJ, Hermes D, Rao RP, denNijs M, and Ojemann JG. Broadband changes in the cortical surface potential track activation of functionally diverse neuronal populations. Neuroimage 85 Pt 2: 711–720, 2014.

20. Crone NE, Miglioretti DL, Gordon B, and Lesser RP. Functional mapping of human sensorimotor cortex with electrocorticographic spectral analysis. II. Event-related synchronization in the gamma band. Brain 121 (Pt 12): 2301–2315, 1998.

21. Miller KJ, Zanos S, Fetz EE, den Nijs M, and Ojemann JG. Decoupling the cortical power spectrum reveals real-time representation of individual finger movements in humans. J Neurosci 29: 3132–3137, 2009.

22. Brucke C, Bock A, Huebl J, Krauss JK, Schonecker T, Schneider GH, Brown P, and Kuhn AA. Thalamic gamma oscillations correlate with reaction time in a Go/noGo task in patients with essential tremor. Neuroimage 75: 36–45, 2013.

23. Chiu SY, Nozile-Firth K, Klassen BT, Adams A, Lee K, Van Gompel JJ, and Hassan A. Ataxia and tolerance after thalamic deep brain stimulation for essential tremor. Parkinsonism Relat Disord 80: 47–53, 2020.

24. Barbe MT, Liebhart L, Runge M, Pauls KA, Wojtecki L, Schnitzler A, Allert N, Fink GR, Sturm V, Maarouf M, and Timmermann L. Deep brain stimulation in the nucleus ventralis intermedius in patients with essential tremor: habituation of tremor suppression. J Neurol 258: 434–439, 2011.

25. Miller KJ, Hermes D, and Staff NP. The current state of electrocorticography-based brain-computer interfaces. Neurosurg Focus 49: E2, 2020.

26. He S, Debarros J, Khawaldeh S, Pogosyan A, Mostofi A, Baig F, Pereira E, Brown P, and Tan H. Closed-loop DBS triggered by real-time movement and tremor decoding based on thalamic LFPs for essential tremor. Annu Int Conf IEEE Eng Med Biol Soc 2020: 3602–3605, 2020.

27. Tan H, Debarros J, He S, Pogosyan A, Aziz TZ, Huang Y, Wang S, Timmermann L, Visser-Vandewalle V, Pedrosa DJ, Green AL, and Brown P. Decoding voluntary movements and postural tremor based on thalamic LFPs as a basis for closed-loop stimulation for essential tremor. Brain Stimul 12: 858–867, 2019.

